# Perinatal and maternal factors associated with Autism Spectrum Disorder

**DOI:** 10.1101/2024.12.22.24319503

**Authors:** Susanna Edlund, Nils Haglund, Carl-Gustaf Bornehag, Chris Gennings, Hannu Kiviranta, Alexander Kolevzon, Christian Lindh, Panu Rantakokko, Abraham Reichenberg, Shanna Swan, Karin Källén

**Author notes:** Corresponding author. (SE).

## Abstract

This study comprehensively examines maternal and perinatal conditions associated with autism spectrum disorder (ASD) in a total population sample with individually confirmed diagnoses from South Sweden. Following thorough review of medical records, 996 cases were ascertained and classified based on level of intellectual disability, ASD severity and family-history of ASD. 10 controls per case were randomly selected from the population (N=9,960). Multiple maternal and perinatal conditions were associated with increased risk for ASD, but associations varied by ASD comorbid conditions. Only high maternal BMI was associated with ASD risk across all ASD sub-groups. Results suggest differences in ASD etiology by comorbid subgroups and highlight potential modifiable risk factors.

## Introduction

Autism spectrum disorder (ASD) is a heterogenous neurodevelopmental disorder associated with a genetic predisposition, and environmental factors. Criteria for ASD as defined by the *Diagnostic and Statistical Manual of Mental Disorders*—5^th^ edition (DSM-5), are persistent deficits in reciprocal social communication as well as restricted and repetitive behaviors including sensory symptoms. Symptoms and severity vary from one individual with ASD to another and an assessment scale based on level of support needed for daily functioning is used to specify severity[1].

ASD is a relatively common diagnosis among children worldwide with an estimated prevalence in Sweden of 1-2% [2]. According to recent large-scale epidemiological studies a 2–5:1 male predominance is found [3,4].

Comorbidity is common and many individuals with ASD face an increased risk of physical and psychological health problems [5,6]. Swedish national clinical guidelines assume that approximately 20-25% of individuals diagnosed with ASD also have an intellectual disability (ID, defined as IQ < 70), making this the most frequent co-occurring condition in ASD [7]. This combination – ASD and ID (ASD+ID), is associated with a worse outcome [8–10].

While it is well-established that ASD is highly heritable [11,12] ranging between 64% to 91% [13], a recent study reported widely different heritability estimates for ASD without ID compared with ASD with ID [14] suggesting potentially different etiological pathways. Furthermore, despite high heritability, the literature suggests that the etiopathogenesis of ASD is complex, and besides genetic factors, environmental factors are also likely to play a role [15,16]. The presumed onset of the development of ASD is during a critical period in pregnancy, when the brain develops and is particularly sensitive to adverse environmental influences [6,17].

Some environmental factors have been repeatedly associated with ASD: prenatal factors such as an advanced maternal age (and paternal age) at birth [9,12,18,19], parity [20], maternal characteristics (e.g., Body Mass Index (BMI), socio-economic status, and country of birth/ethnicity) [21–24], maternal morbidity and/or complications during pregnancy or delivery [25, 26], low/high birth weight [27, 28] and prematurity [29, 30].

The current study combines the advantages of a large cohort and a thorough case evaluation. The role of pre-, peri- and postnatal factors associated with ASD was assessed in a whole and homogenous population of children living in Scania County, including children to mothers who were born in Sweden only. Information regarding factors associated with pregnancy and delivery was obtained using register data with prospectively collected information. Children who were diagnosed with ASD before the age of nine years, were included in the study. All ASD diagnoses were validated by reviewing the complete patient history from birth until the end of the reviewing process in 2019, obtained from medical journals at both the psychiatric clinics and the habilitation clinics. By this thorough medical record review, it was also possible to categorize each case into one of three sub-three sub-groups according to the DSM-V current ASD criteria [1]. Information on familial risks and comorbidities such as ID and/or chromosomal abnormalities was also gathered during this process.

## Material and methods

### The Scania Autism Study Participants

The current study is part of a larger study; The Skania Autism Study (SAS) – a total population study with associated biosamples, established as a resource for environmental ASD research.

The study population and ascertainment process are summarized in Fig 1.

**Fig 1.**
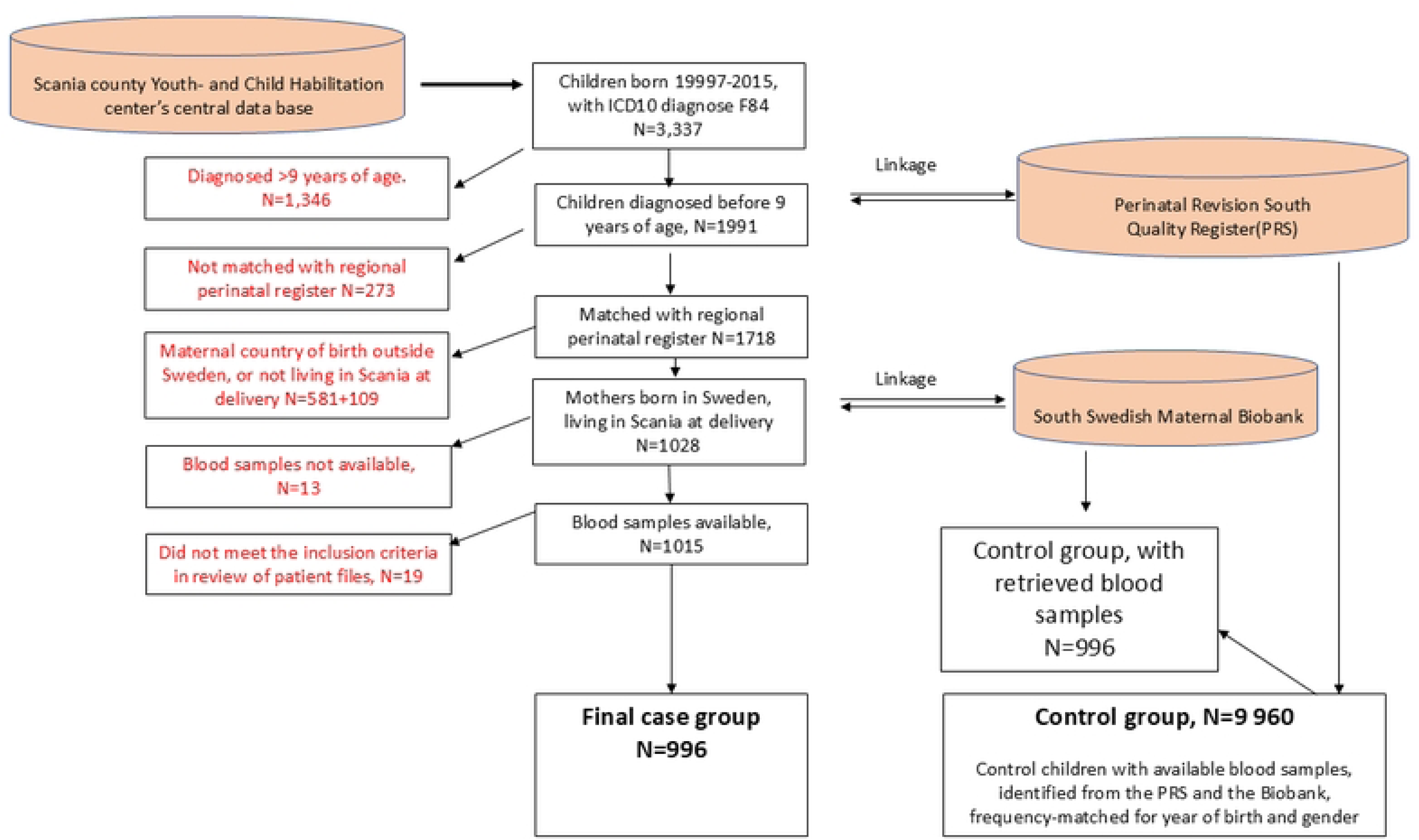
Flow-chart with a visualization of the Autism Spectrum Disorder (ASD) cohort identification and the control selection process.

The central data base for all youth- and habilitation centers in the county of Scania was used to identify all children, born in 1997-2015, with an ICD10-diagnosis of F84 (Pervasive developmental disorders) - including all F84 subcategories (i.e., F84.0 Autistic disorder, F84.5 Asperger’s syndrome and F84.9 Pervasive developmental disorder, unspecified); N=3,337).

Using information on date of the first contact with youth and habilitation services, children who were enrolled (and diagnosed) before the age of nine years were selected (N=1991). Using unique personal ID numbers, the identified children were linked with a regional perinatal quality register (Perinatal Revision South, PRS)[32], which contains data from all obstetric and neonatal units in the southern healthcare region of Sweden, including information on maternal and child characteristics. At the first antenatal visit, information on important background information as maternal age, parity, assisted reproduction, BMI, and chronical diseases is recorded. Information on maternal smoking in early pregnancy, and years of involuntary childlessness is obtained from midwifery interviews. After excluding children whose mothers were not born in Sweden, linkage was possible for 1,028 children, from which maternal blood samples from mid pregnancy could be found for 1,015 children.

Detailed blood samples analyses for several classes of endocrine disruptors including polychlorinated biphenyls (PCBs), perfluorinated chemicals (PFCs), organochlorine pesticides (OCPs), polybrominated diphenyl ethers (PBDEs), and phthalates will be performed at a later stage (as part of the SAS-study).

### Diagnosis confirmation

The medical records of all 1,015 children with available blood samples and clinical data were thoroughly reviewed by an experienced child psychologist (SE) to assess those children who qualified for ASD according to DSM-5 299.00 before the age of nine years. The evaluation was based on best available data, which in most cases included results from the Autism Diagnostic Observation Schedule [33, 34] and the Autism Diagnostic Interview-Revised (ADI-R)[35] (considered to be the “gold standard” assessment measures for ASD). All other assessments scales would be included to give additional information, as would descriptions of patient behaviors during clinical appointments and information given by the patient, parents, schoolteachers and so forth. The medical records from the habilitation clinics were particularly useful when assessing level of severity.

Children with chromosomal anomalies, or confirmed syndromes (e.g., Rett syndrome or Fragile X syndrome) were excluded. The ASD diagnosis was confirmed for 996 children. The level of ASD severity (as specified by DSM-5) was classified as mild, moderate, or severe. A senior child psychologist (NH) reviewed a randomly selected sample of 30 patient records to validate the case review procedure. The inter-observer agreement regarding ASD severity was assessed and was found to be very strong (29 of 30, Kappa index 0.93). A consensus was reached for the one child with discordant ratings after discussion. Information on presence of intellectual disability (ID) was obtained from medical journals. Most cases in the ID-group, had an ID diagnose and had been tested according to standardized clinical assessment of IQ and adaptive functioning. As an exception, a child presenting with obvious cognitive limitations, would sometimes be assigned to the ID-group as suspected but not yet confirmed ID. In those cases, there were among the medical professionals a strong hypothesis of ID; but the child was still waiting for a formal assessment when the recording was done. Also, co-morbidity (Attention-Deficit-Hyperactivity-Disorder (ADHD), cerebral palsy, epilepsy, or hearing- or language-impairment), and heredity was assessed and recorded.

Information on heredity came from the anamnestic information, as obtainable at December 2019, on siblings, parents, grandparents, uncles, aunts, and first cousins. Only confirmed diagnoses were considered (that means, no hearsay information was considered) when classifying familial cases of ASD.

All patient information was stored in a de-identified REDCap electronic data capture tools hosted at Lund University [36].

### Ascertainment of controls

For each child with confirmed ASD, ten controls were randomly selected using the PRS data base, frequency-matched for gender and year of birth, including children to mothers born in Sweden only. Maternal- and child characteristics were retrieved from the PRS data base. One of those controls will be selected for the same type of blood analyses as described above.

### Statistical analyses

Comparisons of gender, intellectual function, reported heredity, or age at diagnosis, respectively, and ASD severity strata were evaluated with Chi-squared test or Mann-Whitney U-test when appropriate. Uni- and multivariable logistic regression analyses were performed to evaluate maternal characteristics, pregnancy- or delivery complications, or newborn characteristics in relation to ASD groups and subgroups. Adjustments were made for maternal age (<20 year, 20-24 years, 25-29 years, 30-34 years, 35-39 years, ≥40 years), parity (primi-multiparity), maternal smoking (no, yes, not known), BMI (<18.5, 18.5-24.9, 25-29.9, ≥30, not known), involuntary childlessness (no, 2-4 years, ≥5 years), assisted reproduction (yes, no). Two-sided p-values below 0.05 were considered statistically significant. All analyses were performed using SPSS version 25.

### Ethics

This study was approved by the Regional Ethical Board in Lund (Dnr 2015/221) with additions by the Swedish Ethical Review Authority (Dnr2020-02348).

This authority also waived patient informed consent.

Data were accessed for research purposes 20220401. The authors had access to patient identity to review medical journals. This process was approved by the ethical authorities.

#### Patient involvement

No patients were involved in defining the research question or outcome measures, nor were they asked to give advice on interpretation of results.

## Results

Table 1 shows the number of ASD cases and controls included in the cohort by matching criteria (year of birth and gender). The overall male-to-female ratio was 4.6:1.

**Table 1.** Children with ASD and controls by matching criteria (year of birth exactly matched, and gender)

|  | Children with ASD<br>N=996 |  | Controls N=9960 |  |
| --- | --- | --- | --- | --- |
|  | n | (%) | n | (%) |
| **Year of birth** |  |  |  |  |
| 1997-2005 | 355 | (35.6) | 3550 | (35.6) |
| 2006-2010 | 438 | (44.0) | 4380 | (44.0) |
| 2011-2015 | 203 | (20.4) | 2030 | (20.4) |
| **Child gender** |  |  |  |  |
| Males | 820 | (82.3) | 8200 | (82.3) |
| Females | 176 | (17.7) | 1760 | (17.7) |

Table 2 shows the ASD severity by age at confirmed diagnosis, sex, intellectual function, and familiarity. There was a significant negative association between age at confirmed diagnosis and ASD severity. The male-to-female ratio did not significantly differ across severity strata. Among children with ASD and intellectual disability, the male-female ratio was 4.0 :1, whereas the corresponding ratio among children with normal intelligence was 4.9:1. The indicated heterogeneity of the male-to-female-ratio across intellectual level strata was not statistically significant (p=0.26) (data not shown).

**Table 2.**
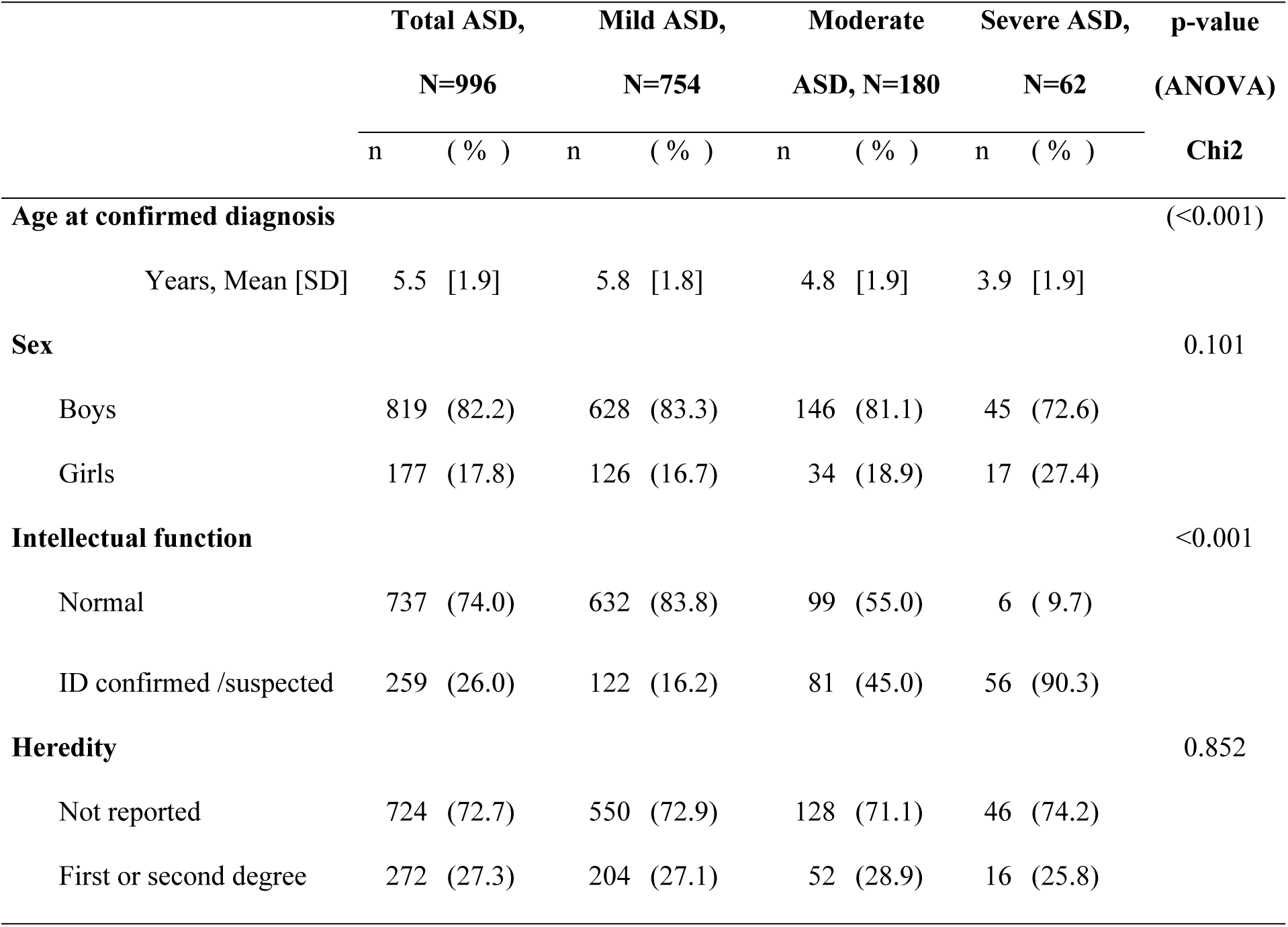
Children with ASD, by ASD severity, presence of intellectual disability (ID), and heredity for ASD.

|  | Total ASD,<br>N=996 |  | Mild ASD,<br>N=754 |  | Moderate<br>ASD, N=180 |  | Severe ASD,<br>N=62 |  | p-value<br>(ANOVA) |
| --- | --- | --- | --- | --- | --- | --- | --- | --- | --- |
|  | n | ( % ) | n | ( % ) | n | ( % ) | n | ( % ) | Chi2 |
| **Age at confirmed diagnosis** |  |  |  |  |  |  |  |  | (<0.001) |
| Years, Mean [SD] | 5.5 | [1.9] | 5.8 | [1.8] | 4.8 | [1.9] | 3.9 | [1.9] |  |
| **Sex** |  |  |  |  |  |  |  |  | 0.101 |
| Boys | 819 | (82.2) | 628 | (83.3) | 146 | (81.1) | 45 | (72.6) |  |
| Girls | 177 | (17.8) | 126 | (16.7) | 34 | (18.9) | 17 | (27.4) |  |
| **Intellectual function** |  |  |  |  |  |  |  |  | <0.001 |
| Normal | 737 | (74.0) | 632 | (83.8) | 99 | (55.0) | 6 | (9.7) |  |
| ID confirmed /suspected | 259 | (26.0) | 122 | (16.2) | 81 | (45.0) | 56 | (90.3) |  |
| **Heredity** |  |  |  |  |  |  |  |  | 0.852 |
| Not reported | 724 | (72.7) | 550 | (72.9) | 128 | (71.1) | 46 | (74.2) |  |
| First or second degree | 272 | (27.3) | 204 | (27.1) | 52 | (28.9) | 16 | (25.8) |  |

A significant association between cognitive level and ASD severity was found. Among children with mild ASD, 16% also had intellectual disability. The corresponding percentage among children with moderate or severe ASD was 45% and 90%, respectively. No association was found between familiarity and level of autism severity.

Tables 3 and 4 present the association between ASD status and maternal characteristics. Table 3 shows the odds ratio (OR) for miscellaneous maternal characteristics for the whole ASD-group compared to controls, whereas Table 4 shows the corresponding ORs by ASD subgroup (ASD severity - mild or moderate/severe, ASD with or without ID, and ASD with or without known heredity, respectively).

**Table 3.** Children with ASD and controls by matching criteria (year of birth and gender), and maternal characteristics.

|  | Children with<br>ASD N=996 |  | Controls N=9960 |  | Odds Ratio |  | Adjusted OR* |  |
| --- | --- | --- | --- | --- | --- | --- | --- | --- |
|  | n | (%) | n | (%) | OR | 95% CI | OR | 95% CI |
| **Maternal age** |  |  |  |  |  |  |  |  |
| <20 | 21 | ( 2.1) | 170 | ( 1.7) | 1.24 | 0.77-2.01 | 0.98 | 0.61-1.57 |
| 20-34 | 775 | (77.8) | 7800 | (78.3) | 1.00 | Reference | 1.0 | Reference |
| 35-39 | 156 | (15.7) | 1682 | (16.9) | 0.91 | 0.75-1.10 | 0.97 | 0.80-1.16 |
| 40+ | 44 | ( 4.4) | 308 | ( 3.1) | 1.36 | 0.96-1.93 | **1.46** | **1.05-2.02** |
| **Parity** |  |  |  |  |  |  |  |  |
| 1 | 518 | (52.0) | 4765 | (47.8) | **1.21** | **1.06-1.39** | **1.23** | **1.07-1.41** |
| 2+ | 478 | (48.0) | 5195 | (52.2) | 1.0 | Reference | 1.0 | Reference |
| **Maternal smoking** |  |  |  |  |  |  |  |  |
| Yes | 138 | (13.9) | 940 | ( 9.4) | **1.55** | **1.27-1.89** | **1.49** | **1.22-1.82** |
| No | 826 | (82.9) | 8748 | (87.8) | 1.00 | Reference | 1.0 | Reference |
| *Not known* | 32 | ( 3.2) | 272 | ( 2.7) | 0.86 | 0.46-1.60 | 1.01 | 0.68-1.50 |
| **Maternal BMI** |  |  |  |  |  |  |  |  |
| <18.5 | 21 | ( 2.1) | 184 | ( 1.8) | 1.55 | 0.98-2.46 | 1.53 | 0.96-2.43 |
| 18.5-24.9 | 434 | (43.6) | 5404 | (54.3) | 1.0 | Reference | 1.0 | Reference |
| 25-29.9 | 280 | (28.1) | 2524 | (25.3) | **1.40** | **1.20-1.64** | **1.41** | **1.20-1.65** |
| 30+ | 178 | (17.9) | 1269 | (12.7) | **1.79** | **1.49-2.16** | **1.76** | **1.46-2.12** |
| *Not known* | 83 | ( 8.3) | 579 | ( 5.8) | **1.78** | **1.39-2.28** | **1.78** | **1.36-2.31** |
| **Involuntary childlessness** |  |  |  |  |  |  |  |  |
| No (<2 years) | 938 | (94.2) | 9438 | (94.8) | 1.00 | Reference | 1.0 | Reference |
| 2-4 years | 44 | ( 4.4) | 416 | ( 4.2) | 1.08 | 0.78-1.50 | 1.02 | 0.71-1.45 |
| 5 years or more | 14 | ( 1.4) | 106 | ( 1.1) | 1.14 | 0.61-2.12 | 1.14 | 0.61-2.10 |
| **Assisted reproduction** |  |  |  |  |  |  |  |  |
| Yes | 30 | ( 3.0) | 278 | ( 2.8) | 1.08 | 0.74-1.58 | 1.02 | 0.64-1.60 |
| No | 966 | (97.0) | 9682 | (97.2) | 1.0 | Reference | 1.0 | Reference |

**Table 4.**
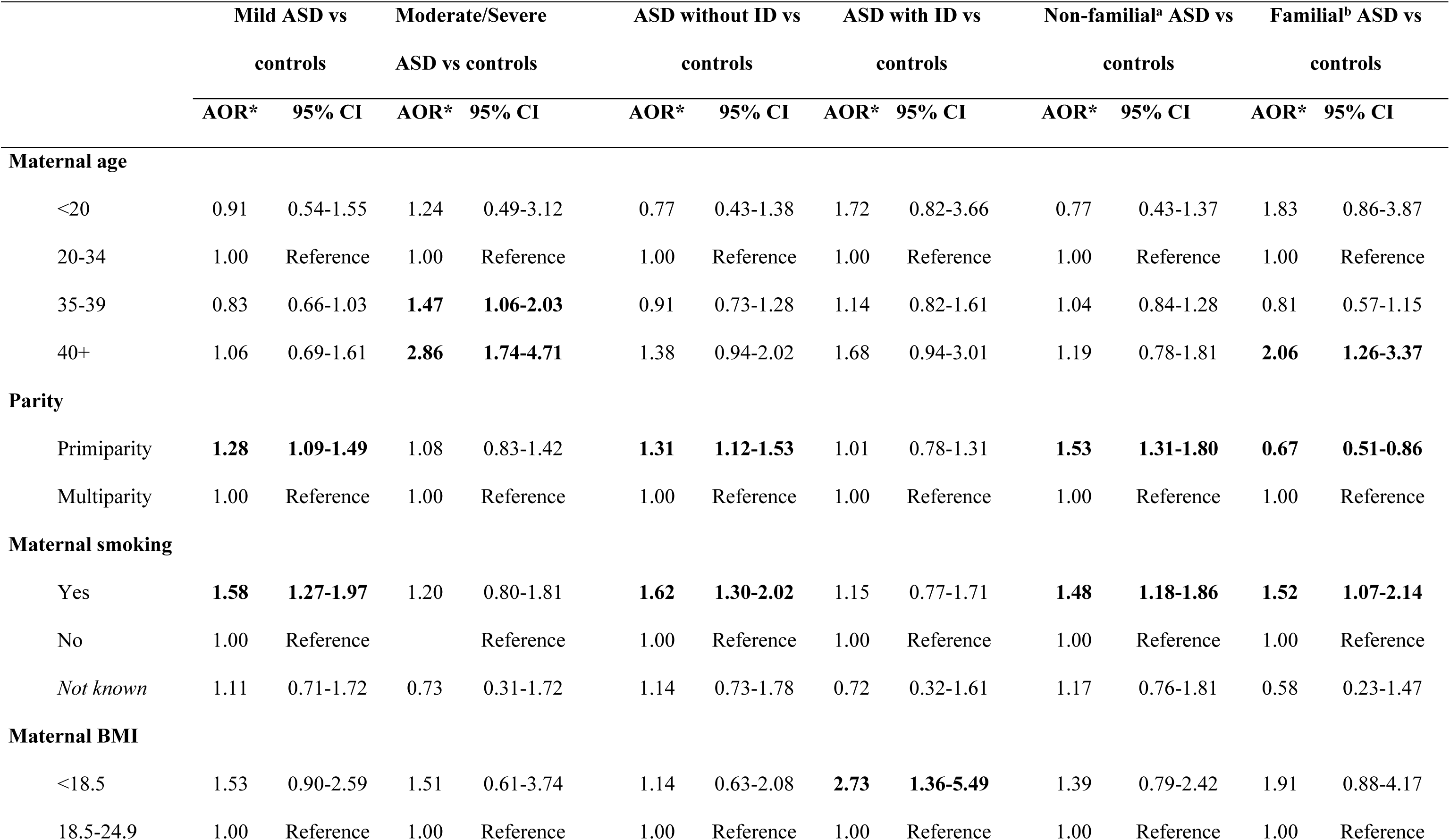
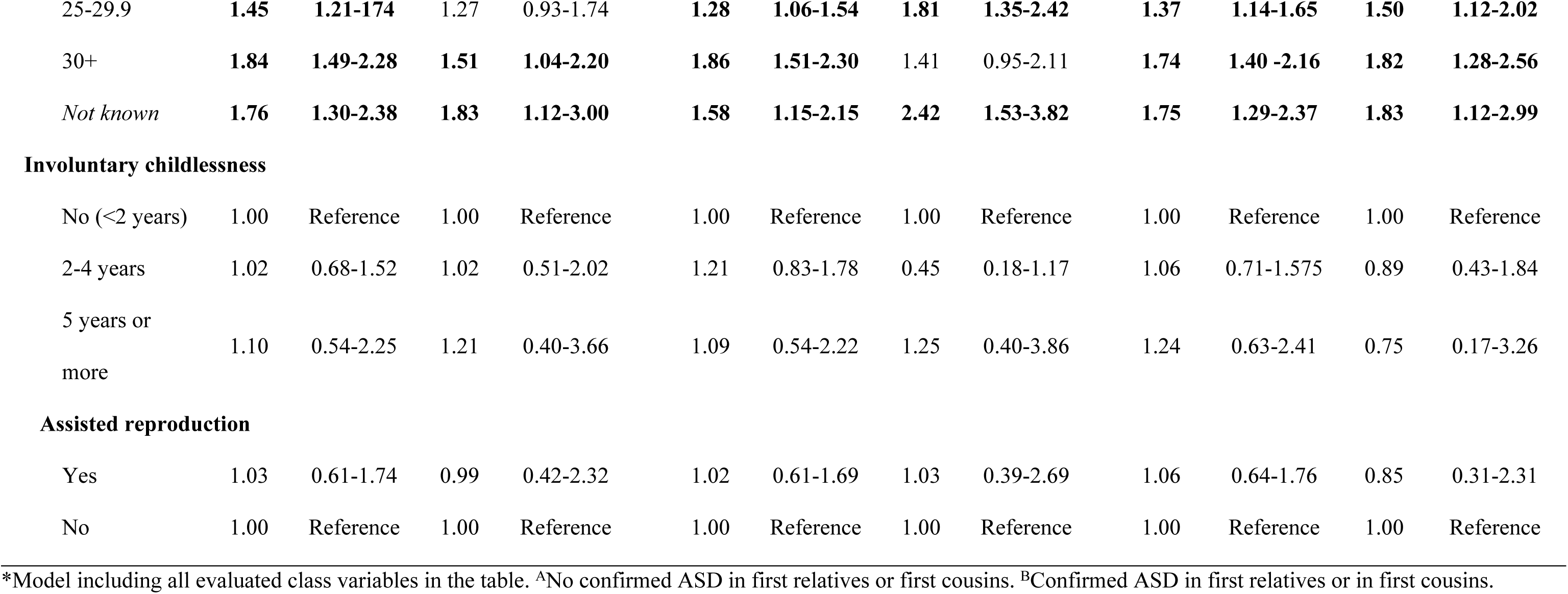
Autism severity, presence of intellectual disability, and heredity, respectively, by maternal characteristics.

After adjustments, significantly increased ORs for offspring ASD (all groups) were found for maternal age 40 or older (Table 3). The association between ASD and advanced maternal age was evident only for moderate/severe ASD, and for familial ASD (Table 4). Children with ASD were more often born to primiparous women than were control children. This was true for mild ASD and/or ASD without ID.

Maternal smoking was a significant factor associated with ASD, for all ASD subgroups. Among the evaluated maternal factors, maternal overweight (BMI 25-29.9), obesity (BMI ≥ 30), or no reported maternal BMI, constituted the strongest factors associated with ASD. Furthermore, a strong and significant association between increasing BMI and ASD was found for almost all ASD subgroups. Maternal underweight (BMI <18.5) was associated with ASD+ID but was not associated with any other ASD subgroup compared to controls. A u-shaped association between ASD and BMI was indicated, but far from statistically significant (p>0.9). No increased association with ASD was seen among women who had experienced involuntary childlessness or had underwent assisted reproductive treatments.

Pregnancy complications and infant characteristics among children with ASD and controls are presented in Table 5 and Table 6. An association between “any pregnancy complication” and ASD was primarily indicated but was attenuated after adjustments for maternal characteristics. Gestational diabetes was the only individual pregnancy complication for which an association with ASD was initially found, but the association was no longer statistically significant when adjusted for BMI. Gestational diabetes was associated with moderate/severe ASD and with ASD-ID even after adjustments for maternal characteristics (Table 6).

**Table 5.**
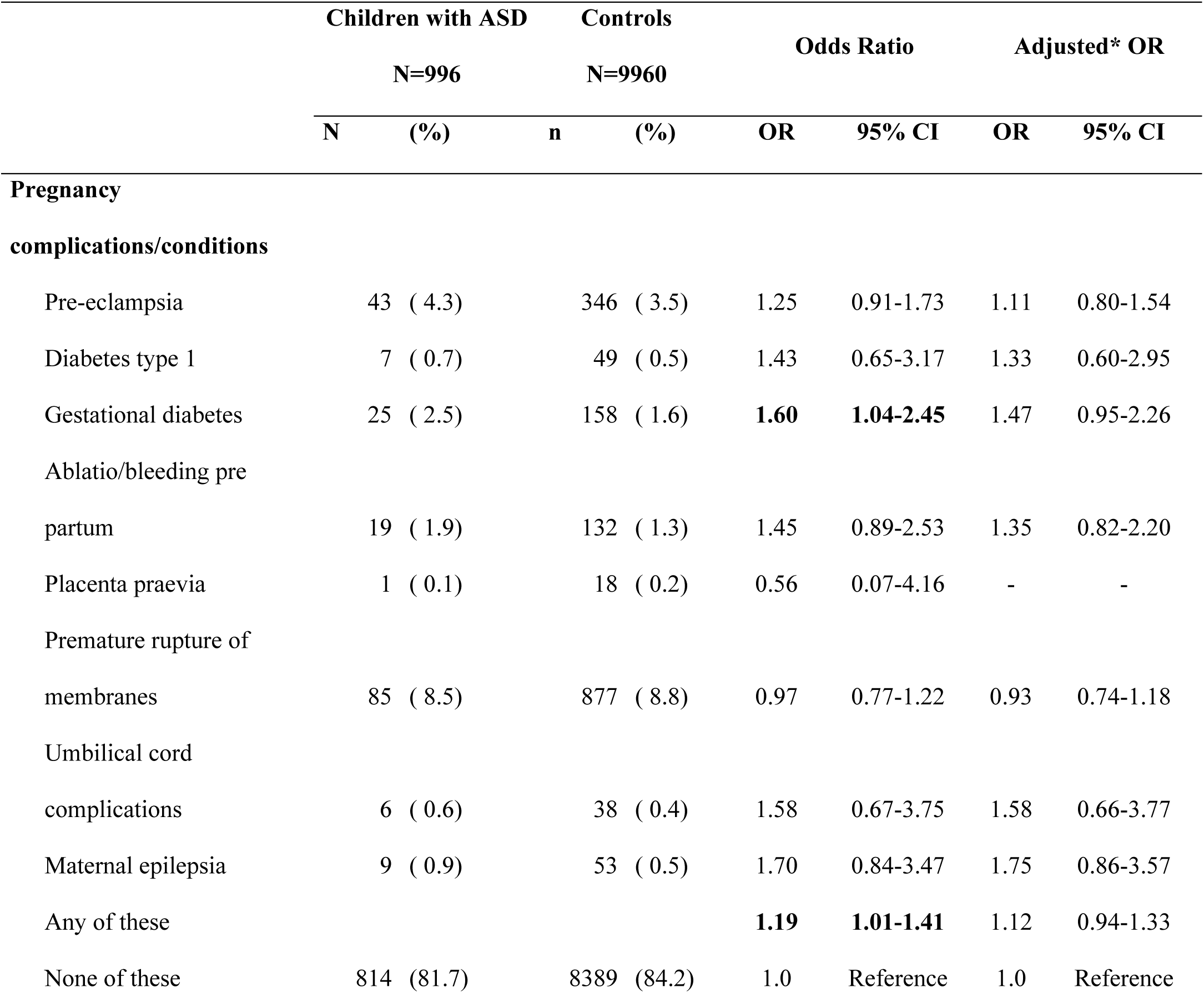
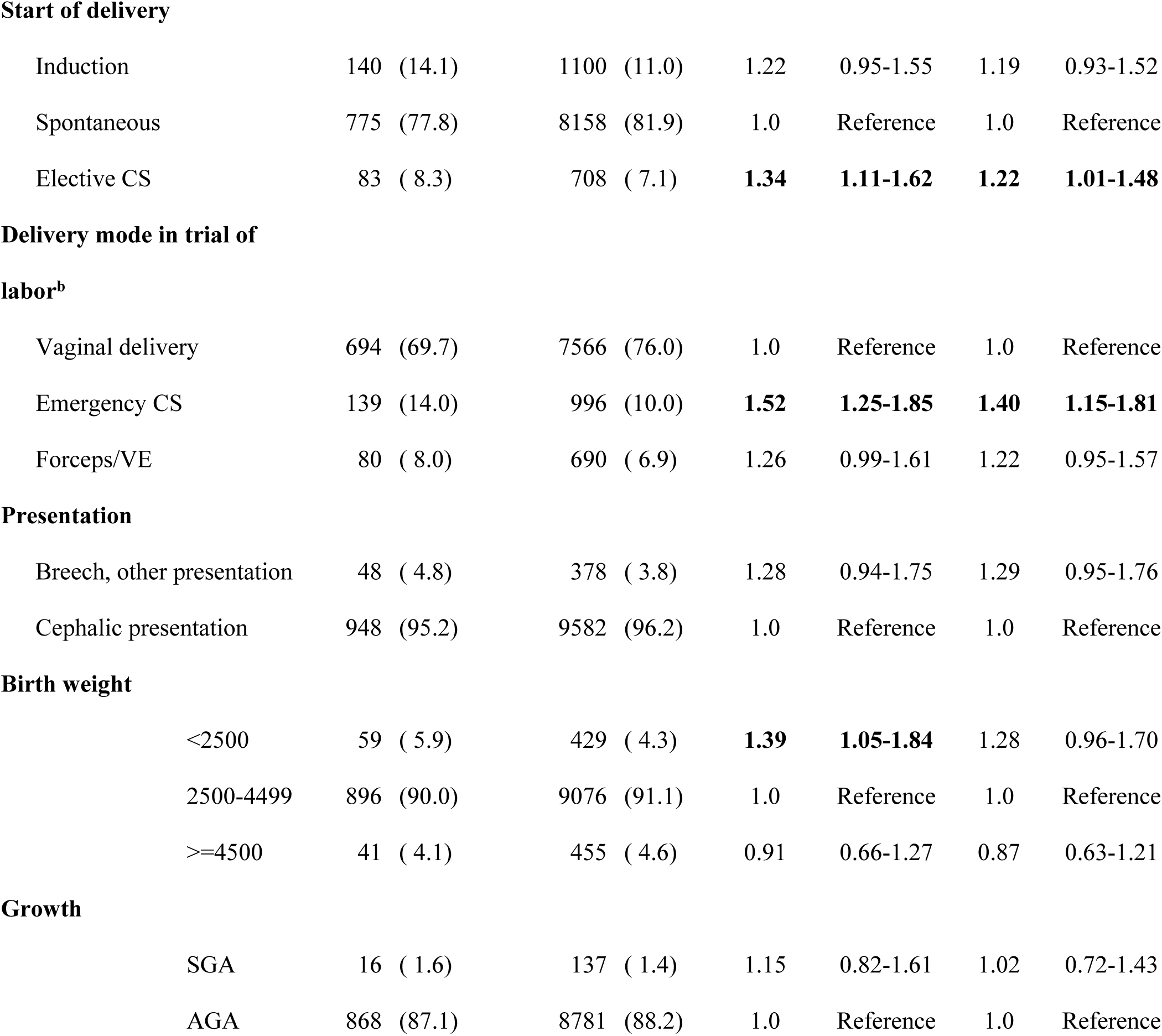
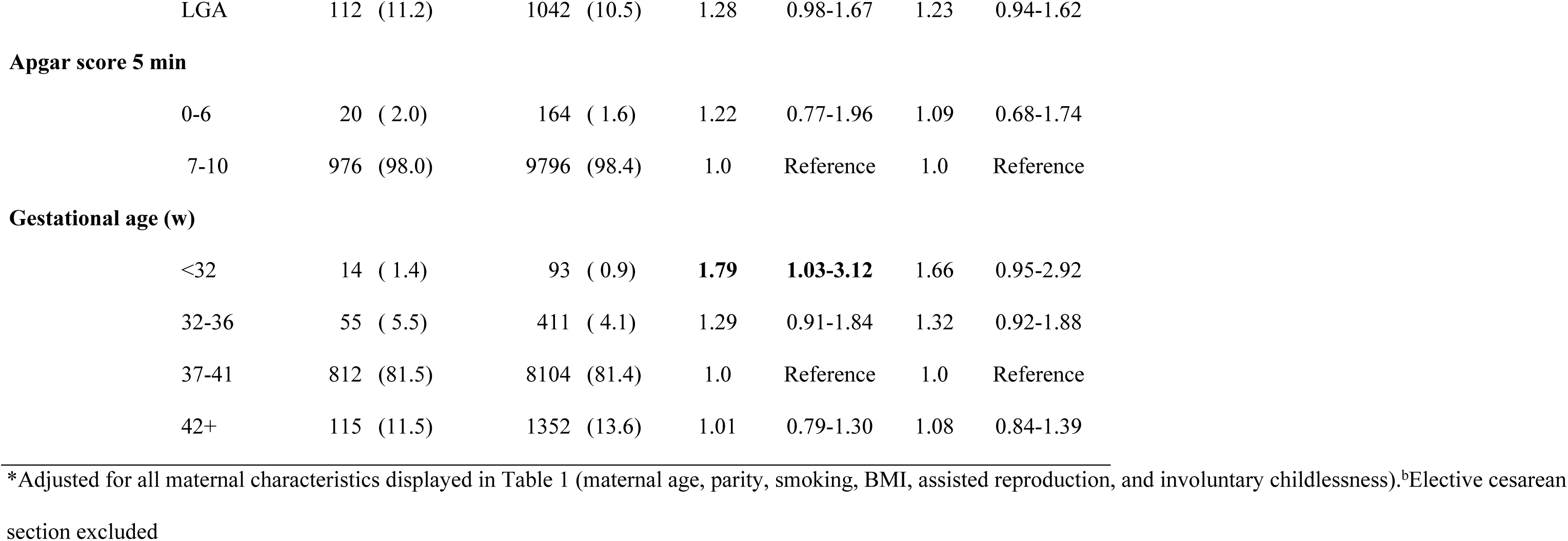
Children with ASD and controls, by presence of pregnancy, delivery complications, and infant characteristics.

|  | Children with ASD |  | Controls |  | Odds Ratio |  | Adjusted* OR |  |
| --- | --- | --- | --- | --- | --- | --- | --- | --- |
|  | N=996 |  | N=9960 |  |  |  |  |  |
|  | N | (%) | n | (%) | OR | 95% CI | OR | 95% CI |
| **Pregnancy** |  |  |  |  |  |  |  |  |
| **complications/conditions** |  |  |  |  |  |  |  |  |
| Pre-eclampsia | 43 | ( 4.3) | 346 | ( 3.5) | 1.25 | 0.91-1.73 | 1.11 | 0.80-1.54 |
| Diabetes type 1 | 7 | ( 0.7) | 49 | ( 0.5) | 1.43 | 0.65-3.17 | 1.33 | 0.60-2.95 |
| Gestational diabetes | 25 | ( 2.5) | 158 | ( 1.6) | **1.60** | **1.04-2.45** | 1.47 | 0.95-2.26 |
| Ablatio/bleeding pre |  |  |  |  |  |  |  |  |
| partum | 19 | ( 1.9) | 132 | ( 1.3) | 1.45 | 0.89-2.53 | 1.35 | 0.82-2.20 |
| Placenta praevia | 1 | ( 0.1) | 18 | ( 0.2) | 0.56 | 0.07-4.16 | - | - |
| Premature rupture of |  |  |  |  |  |  |  |  |
| membranes | 85 | ( 8.5) | 877 | ( 8.8) | 0.97 | 0.77-1.22 | 0.93 | 0.74-1.18 |
| Umbilical cord |  |  |  |  |  |  |  |  |
| complications | 6 | ( 0.6) | 38 | ( 0.4) | 1.58 | 0.67-3.75 | 1.58 | 0.66-3.77 |
| Maternal epilepsia | 9 | ( 0.9) | 53 | ( 0.5) | 1.70 | 0.84-3.47 | 1.75 | 0.86-3.57 |
| Any of these |  |  |  |  | **1.19** | **1.01-1.41** | 1.12 | 0.94-1.33 |
| None of these | 814 | (81.7) | 8389 | (84.2) | 1.0 | Reference | 1.0 | Reference |

**Start of delivery**
|  |  |  |  |  |  |  |
| --- | --- | --- | --- | --- | --- | --- |
| Induction | 140 (14.1) | 1100 (11.0) | 1.22 | 0.95-1.55 | 1.19 | 0.93-1.52 |
| Spontaneous | 775 (77.8) | 8158 (81.9) | 1.0 | Reference | 1.0 | Reference |
| Elective CS | 83 ( 8.3) | 708 ( 7.1) | **1.34** | **1.11-1.62** | **1.22** | **1.01-1.48** |

**Delivery mode in trial of****labor<sup>b</sup>**
|  |  |  |  |  |  |  |
| --- | --- | --- | --- | --- | --- | --- |
| Vaginal delivery | 694 (69.7) | 7566 (76.0) | 1.0 | Reference | 1.0 | Reference |
| Emergency CS | 139 (14.0) | 996 (10.0) | **1.52** | **1.25-1.85** | **1.40** | **1.15-1.81** |
| Forceps/VE | 80 ( 8.0) | 690 ( 6.9) | 1.26 | 0.99-1.61 | 1.22 | 0.95-1.57 |

**Presentation**
|  |  |  |  |  |  |  |
| --- | --- | --- | --- | --- | --- | --- |
| Breech, other presentation | 48 ( 4.8) | 378 ( 3.8) | 1.28 | 0.94-1.75 | 1.29 | 0.95-1.76 |
| Cephalic presentation | 948 (95.2) | 9582 (96.2) | 1.0 | Reference | 1.0 | Reference |

**Birth weight**
|  |  |  |  |  |  |  |
| --- | --- | --- | --- | --- | --- | --- |
| <2500 | 59 ( 5.9) | 429 ( 4.3) | **1.39** | **1.05-1.84** | 1.28 | 0.96-1.70 |
| 2500-4499 | 896 (90.0) | 9076 (91.1) | 1.0 | Reference | 1.0 | Reference |
| ≥4500 | 41 ( 4.1) | 455 ( 4.6) | 0.91 | 0.66-1.27 | 0.87 | 0.63-1.21 |

**Growth**
|  |  |  |  |  |  |  |
| --- | --- | --- | --- | --- | --- | --- |
| SGA | 16 ( 1.6) | 137 ( 1.4) | 1.15 | 0.82-1.61 | 1.02 | 0.72-1.43 |
| AGA | 868 (87.1) | 8781 (88.2) | 1.0 | Reference | 1.0 | Reference |
| LGA | 112 (11.2) | 1042 (10.5) | 1.28 | 0.98-1.67 | 1.23 | 0.94-1.62 |
| **Apgar score 5 min** |  |  |  |  |  |  |
| 0-6 | 20 (2.0) | 164 (1.6) | 1.22 | 0.77-1.96 | 1.09 | 0.68-1.74 |
| 7-10 | 976 (98.0) | 9796 (98.4) | 1.0 | Reference | 1.0 | Reference |
| **Gestational age (w)** |  |  |  |  |  |  |
| <32 | 14 (1.4) | 93 (0.9) | **1.79** | **1.03-3.12** | 1.66 | 0.95-2.92 |
| 32-36 | 55 (5.5) | 411 (4.1) | 1.29 | 0.91-1.84 | 1.32 | 0.92-1.88 |
| 37-41 | 812 (81.5) | 8104 (81.4) | 1.0 | Reference | 1.0 | Reference |
| 42+ | 115 (11.5) | 1352 (13.6) | 1.01 | 0.79-1.30 | 1.08 | 0.84-1.39 |

\*Adjusted for all maternal characteristics displayed in Table 1 (maternal age, parity, smoking, BMI, assisted reproduction, and involuntary childlessness).<sup>b</sup>Elective cesarean section excluded

**Table 6.**
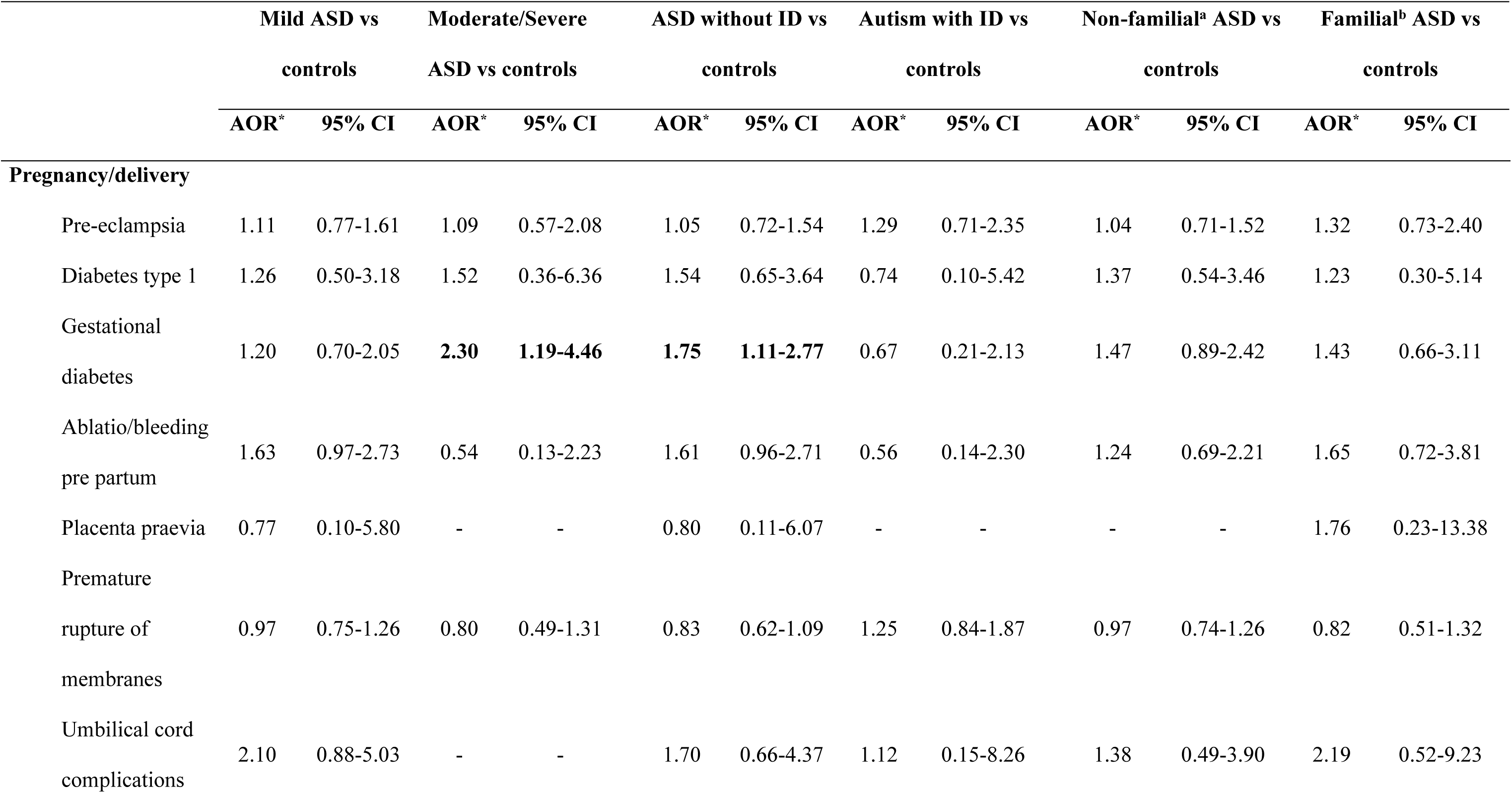
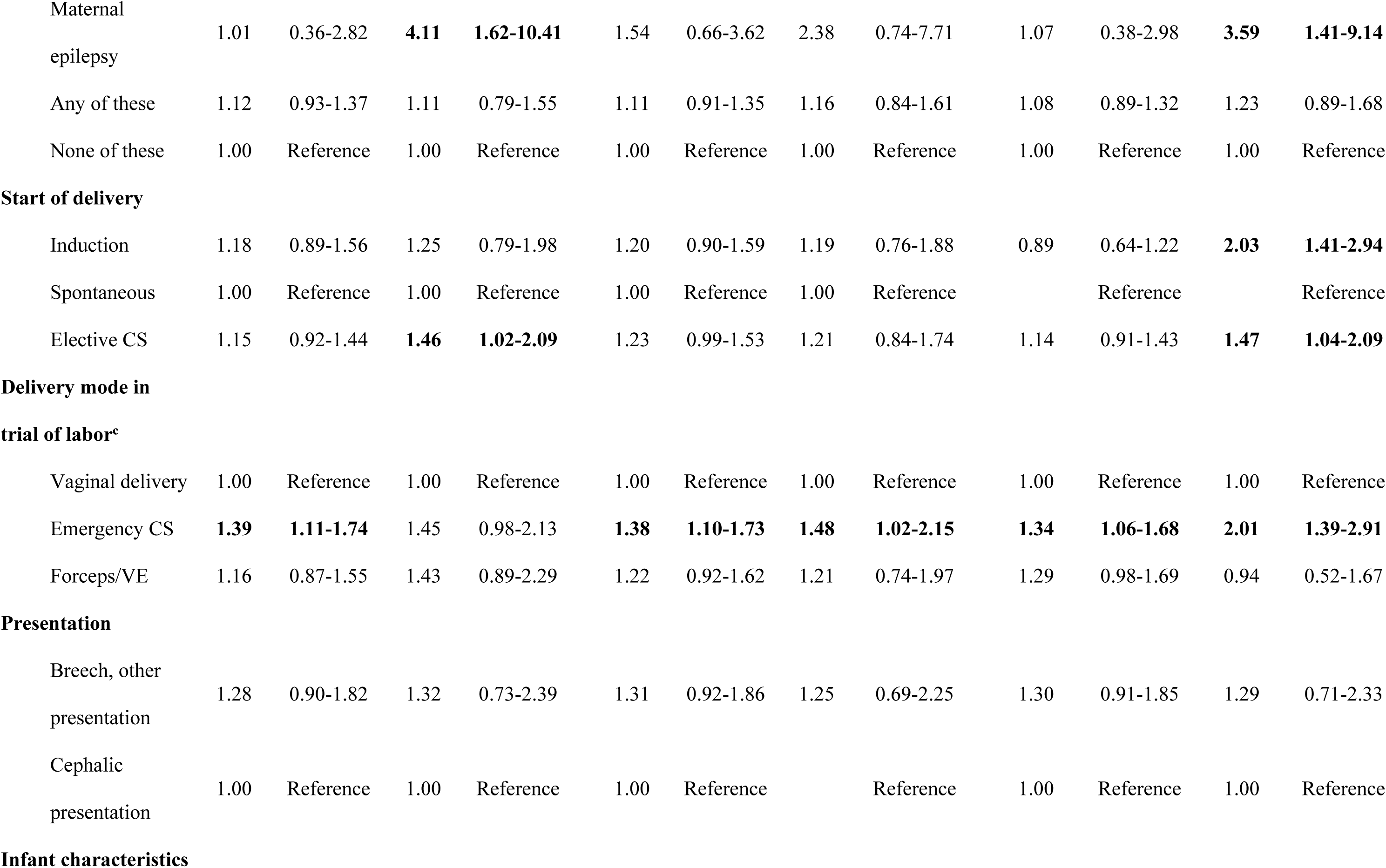
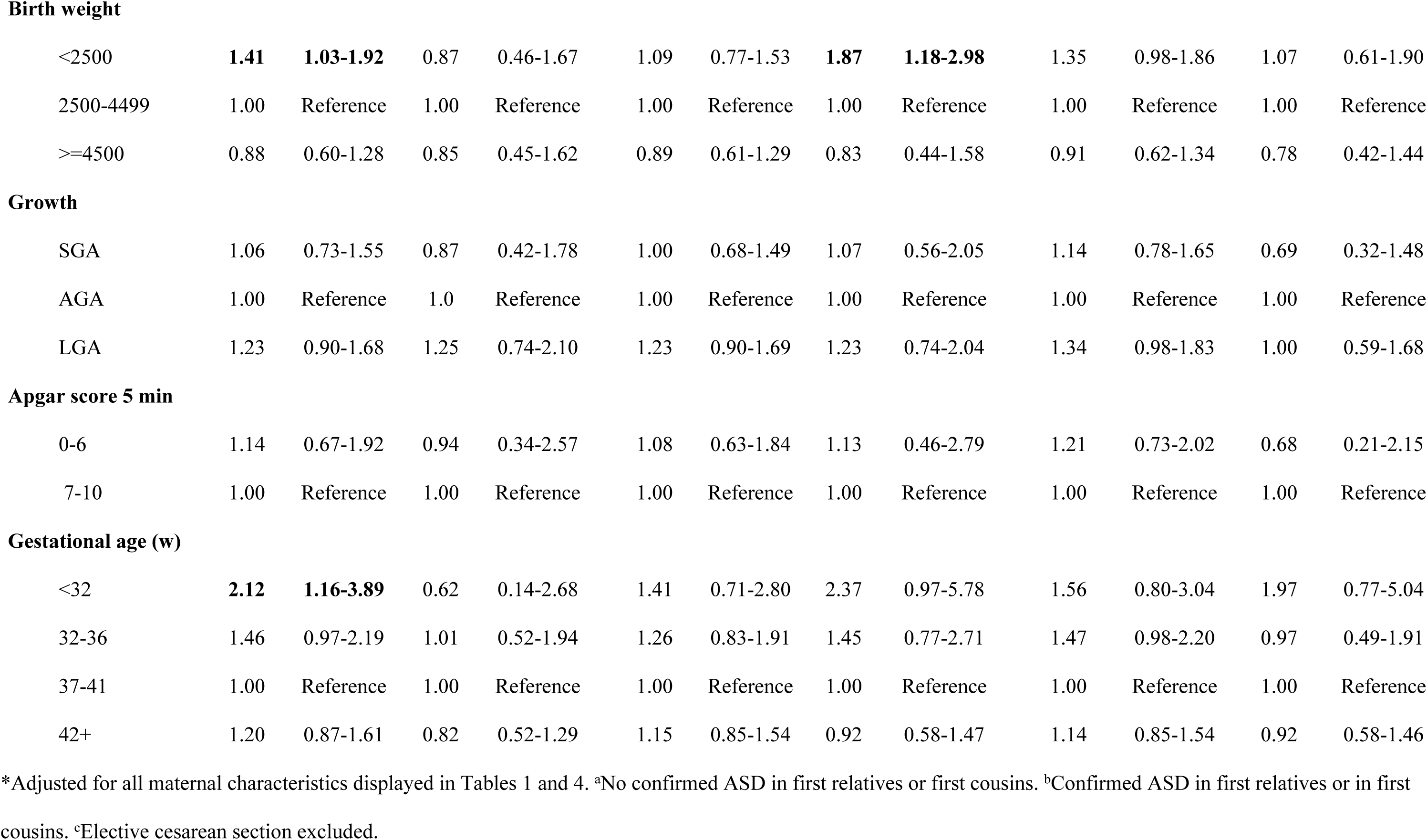
Autism Spectrum Disorder (ASD) severity and presence of Intellectual Disability (ID) respectively, by pregnancy complications, delivery mode, and infant.

Maternal epilepsy was associated with moderate/severe ASD, and familial ASD, respectively. Children with ASD were more often delivered by planned or emergency cesarean section (CS) (Table 5). Emergency CS was associated with all ASD subgroups, whereas elective CS was associated with moderate/severe ASD and familial ASD only (Table 6). An association between low birth weight and/or very preterm birth (<32 weeks) was found but was not statistically significant after adjustment. For mild ASD, a significant association with very preterm birth, and low birth weight (<2500g), respectively, was evident also after adjustments. No association between low Apgar score and ASD was observed.

## Discussion

Using a large population-based cohort with validated diagnosis and detailed information on ASD severity, cognitive level, and familial history, linked to a population-based perinatal database, we were able to simultaneously investigate a large variety of maternal and pregnancy related factors associated with ASD and for ASD subgroups.

Maternal smoking, overweight, obesity, elective CS, and emergency CS, respectively, were factors associated with ASD without any substantial heterogeneity among ASD subgroups. Advanced maternal age was associated with moderate/severe ASD and familial ASD but not for other sub-groups. One or more ASD subgroups were associated with gestational diabetes, maternal epilepsy, low birth weight (<2500g), and very preterm birth (<32 weeks) (mild ASD only). Otherwise, few pregnancy complications and perinatal conditions were over-represented in the ASD group.

The most striking finding of the current study was the strong and significantly increased association for ASD in children born to mothers with overweight or obesity. The association was present in all subgroups, no matter which ASD subtype or status of ID. Adjustments for other maternal characteristics only marginally altered the odds ratio estimates. Pregnancy obesity has been linked to several endocrine disruptions, such as increased systemic inflammation / adiposity-induced inflammation, insulin resistance, affected endocrine responses, and in utero steroid dysregulation [37, 38]. Disturbances like these, in the maternal-fetal-placental unit, seem to increase the association of suboptimal neurological effects and several studies show associations between pre-pregnancy obesity and neurodevelopmental disorders (24,39,40]. In one major meta-analytic study, the association between overweight and ASD was eliminated when adjusting for familial confounding [41]. In our study, the increased association remained in the familial group, and the magnitude of the association between BMI and ASD did not differ between cases with or without heredity for ASD. Underweight was associated with one of the subgroups (ASD+ID). The association between both overweight/obesity and underweight has previously been reported in two large population-based studies, where the authors conclude that extremes in maternal weight increases the association with ASD [42, 43]. In the current study, no information on weight gain during pregnancy was available.

Exposure to tobacco smoke is believed to disrupt the fetal brain development in several ways; including direct toxicity when smoking occurs and indirect negative effects on placental function [44]. It has been suggested [45] that smoking during pregnancy is stronger linked to other mental health conditions rather than to ASD. We did though, find a significant association between maternal smoking and ASD - most pronounced for mild ASD and/or ASD – ID. These results are in line with von Ehrenstein et al.,[46] who, when investigating the smoking habits among mothers of 11 722 ASD cases, found a significant association between heavy smoking and ASD, and a higher estimate for mild than for moderate/severe ASD. The quoted study was not included in a meta-analysis published in 2015 in which no evidence to support a measurable association between maternal prenatal smoking and ASD in offspring was found [47]. A recent meta-analysis containing 72 extant cohorts in the Environmental Influences on Child Health Outcomes consortium in US, found initially no associated risk of ASD when studying effects on maternal smoking [44]. However, after excluding studies with small cell sizes and studies containing cases of preterm birth, a significant association between maternal smoking and ASD was found (OR 1.44). The researchers hypothesize that the third trimester could be a critical window during which the fetus would be more sensitive to certain air borne exposures [48, 49]. In our study, information on smoking were obtained during the first trimester. Mothers reporting to smoke would then be strongly advised to quit smoking and probably several would follow this advice. Information on smoking in mid-pregnancy is available in the Swedish Medical Birth register, but not reliable due to missing data. If maternal smoking in late pregnancy is important for the development of ASD and a large percentage of the women who smoke in early pregnancy would have quitted smoking during pregnancy, the association between maternal smoking and ASD may be underestimated in our study. To what extent the association between smoking and ASD is causal or if the association is confounded by maternal characteristics is not fully known.

Children with ASD were significantly more likely to be delivered by emergency CS than controls, suggesting that although no specific pregnancy complications were recorded, deliveries with children with ASD were more likely to have been exposed to hazardous conditions during late pregnancy or during delivery compared to controls. However, despite the high rate of emergency CS among children with ASD, no association between low Apgar score at five minutes and ASD was found. Elective CS was also more common among children with ASD than among controls. In the absence of pregnancy complications causing indications for planned CS, the association could perhaps be explained by maternal psychological factors in the ASD group such as an increased prevalence of anxiety among mothers of children with ASD. The detailed control from potential confounding factors in present study adds to previous work [25,50], which also reported significant associations between ASD and both emergency and planned CS.

Multiple studies have reported an association between advanced maternal age and ASD [23,51,52]. One hypothesis is that increased maternal age also increases the risk of chromosomal abnormalities or unstable trinucleotide repeats [51], alternatively advanced maternal age adds to the risk of pregnancy/birth complications leading to adverse birth outcomes [23,52]. In the current study, a significant association between advanced maternal age and ASD was found for moderate/severe ASD and familial ASD, even though children with chromosomal abnormalities were excluded. Furthermore, the association between advanced maternal age and ASD was not due to pregnancy complications, delivery mode, or prematurity. Thus, other factors should account for the observed association. One possibility is that women with mild ASD traits may be more likely to find their partners later in life. This explanation may be supported by the finding that advanced maternal age was more associated with ASD with reported heredity than ASD without reported heredity. Also, paternal age may be an important confounder. Information of paternal age was not collected. Based on statistical information on age differences in heterogenic relationships in Sweden, the man is younger than the woman in only 20 % cases [53]. Most children in the group advanced maternal age, would then probably also have older fathers.

The estimates for familial ASD vary substantially between studies ranging from 38% to 90% [11,54–56]. The difficulty to reach a consensus concerning the degree of familiarity depends, among other things, on the estimation methods, reported ASD prevalence, and inclusion criteria. In studies where heredity is estimated using the ASD concordance differences between monozygotic and dizygotic twins, a higher degree of heredity is reported when ASD is defined using clinically confirmed diagnostic criteria [13,57]. In the current study, the goal was not to estimate the heritability in ASD, but we conclude that 27% of the children with ASD had a first or second degree (including cousins) relative with a confirmed diagnosis of ASD, and the frequency did not differ between ASD severity groups.

Primiparity is associated with pregnancy and delivery complications and has frequently been associated with an increased association of several adverse neurodevelopmental outcomes, including ASD [51,58,59]. In the present study, primiparity was associated with ASD, in particularly among children with mild ASD and ASD – ID, which has previously been reported [60,61].

In the multivariable analyses, no associations between pregnancy complications and ASD were detected when analyzing the entire cohort. Gestational diabetes was associated with ASD in the crude analysis, but when maternal BMI was adjusted for, no significant association remained. An association between gestational diabetes and ASD has previously been reported. In a recent meta-analysis, Wan et al., [62] detected high heterogeneity among studies, and they also detected evidence of publication bias. When they selected only high-quality case control studies, a significant association between ASD and gestational diabetes was detected.

Epilepsy has frequently been shown to be associated with an increased risk of maternal and offspring mortality and morbidity, including an increased association with ASD in offspring [63]. Although no association between maternal epilepsy and ASD was found in the entire cohort in the current study, sub-group analyses revealed significant associations between maternal epilepsy and both hereditary- and moderate/severe ASD. Medications used for epilepsy, especially valproic acid, are potential human teratogens. They are also suspected to cause delayed neurodevelopment [38,64,65]. Whether or not it is the medication or maternal epilepsy per se that causes this association is not fully known. Maternal use of valproate during pregnancy was significantly associated with ASD in the offspring, even after adjusting for maternal epilepsy [66].

Several recent meta-analyses [23,30,67] found evidence for an association between ASD and preterm birth, while another large, albeit older, review did not detect such an association even though a large heterogeneity among studies was identified [68]. In the current study, a significant association between mild ASD and very preterm birth was found (with subsequent low birth weight) – otherwise no association between gestational duration and ASD was found.

Even though we have not been able to categorize all children with ASD according to genetic predisposition for ASD, we have used the material available to divide the children into probably high genetic risk and probably low genetic risk. We wanted to explore if there was a heterogeneity within the risk factor spectrum among the familial group and the non-familial. A strong, external risk factor would perhaps be hidden in a group who would have developed ASD, regardless of factors present in pregnancy or delivery. Therefore, the material was divided into familial and non-familial cases. Our results indicated a stronger association between maternal epilepsy and ASD in the offspring among familial than non-familial cases, suggesting that in our material, maternal epilepsy is more likely to be a part of the genetic spectrum related to ASD. In the case of gestational diabetes, no heterogeneity between familial and non-familial cases was indicated. Thus, gestational diabetes in our material, could be interpreted to be a true primary risk factor for ASD in the offspring. The same may be true for delivery complications that made it necessary to perform an emergency cesarean section. The AORs for maternal smoking and high BMI were similar among familial and non-familial cases, which could imply that maternal smoking and high maternal BMI are true risk factors for ASD in the children.

### Strengths and limitations

The current study is population-based and includes all children with ASD who were diagnosed and treated at a center for youth- and child habilitation in the county of Scania. Each case was thoroughly scrutinized and linked to a high-quality perinatal database. Thus, the current study combines the advantages of a large population-based registry study with the case ascertainment precision of a case-control study. The population was homogenous since we constricted the data to include children to mothers born in Sweden only. A wide array of possible associated factors with ASD were evaluated, including both pre-, peri- and post-natal events. However, we did not have access to parental socio-economic status and educational level, which may be considered as a limitation of the current study. Obesity is a well-known marker of low social class and/or low educational level [69]. Thus, if a strong association exists between ASD and parental low social status, the association between high BMI and ASD found in the current study could be due to confounding from socio-economic factors - at least to some extent. Furthermore, we had no information on paternal characteristics, such as smoking, age, or BMI.

## Conclusion

Several associated factors for ASD were elucidated in this large, population-based epidemiological study, and we replicate results from previous studies. In particular, the association between overweight/obesity or maternal smoking and ASD, respectively, was present among all (or nearly all) ASD subgroups. Our main findings are especially important given that both these associated factors are modifiable and may be significantly reduced through education and medical intervention.

## Data Availability

The register data are not publicly available due to privacy protection, including General Data Protection Regulations (GDPR). Access to Swedish register resoruces is only granted after ethical review by appropriate authorities. Requests should be directed to.

